# Effectiveness of dexamethasone as an adjuvant to intrathecal bupivacaine versus bupivacaine alone in spinal anesthesia among orthopedic surgery patients at KCMC referral hospital, northern Tanzania

**DOI:** 10.64898/2026.05.18.26353515

**Authors:** Kevin Fidelis, Edwin Shewiyo, William Nkenguye, Baraka Kawiche, Gregory Goodluck, Lyidia V Masika, Anna Dohho, Maynard Mekere, Victoria Adonicam, Fredson Mwiga, Happyness sway, Alex Lwiza, Shuweikha S Mohammed, Brian Vaughan, Nyasatu Chamba

## Abstract

**Background:** Orthopedic surgeries are associated with significant intraoperative and postoperative pain, necessitating effective anesthesia strategies. Spinal anesthesia is commonly used for lower limb procedures due to its rapid onset and reliability; however, its limited duration may compromise prolonged surgical procedures and early postoperative pain control. Adjuvants such as dexamethasone have been explored to enhance and prolong the effects of local anesthetics. While evidence supports its efficacy, data from low-resource settings remain limited.

**Objective:** To assess the effect of intrathecal dexamethasone as an adjuvant to bupivacaine on sensory block duration, time to first postoperative analgesia, and postoperative pain in patients undergoing lower limb orthopedic surgery at KCMC.

**Methodology:** A randomized, double-blind controlled trial was conducted among 96 adult patients undergoing elective lower limb orthopedic surgery under spinal anesthesia. Participants were allocated using a computer-generated randomization sequence to receive either bupivacaine 15 mg with dexamethasone 4 mg (intervention group) or bupivacaine 15 mg with 1 ml normal saline (control group). Outcomes included sensory and motor block duration, time to first postoperative analgesia, and postoperative pain scores.

**Results:** The dexamethasone group demonstrated a significantly prolonged sensory block duration (231 ± 6 vs. 156 ± 9 minutes; mean difference 75.11 minutes, 95% CI: 71.92– 78.29; p < 0.001) and delayed time to first postoperative analgesia (252 ± 7 vs. 181 ± 7 minutes; mean difference 71.89 minutes, 95% CI: 68.91–74.86; p < 0.001). Motor block duration was also significantly longer (184 ± 7 vs. 130 ± 5 minutes; mean difference 53.42 minutes, 95% CI: 50.99–55.85; p < 0.001). Postoperative pain scores were significantly lower at 1 hour (mean difference −1.29 points, 95% CI: −1.52 to −1.05; p < 0.001) and at 2 hours (mean difference −1.97 points, 95% CI: −2.21 to −1.73; p < 0.001). Intraoperative opioid and benzodiazepine use were significantly reduced in the intervention group.

**Conclusion:** The addition of intrathecal dexamethasone to bupivacaine significantly enhances sensory block duration, delays postoperative analgesia need, and improves early pain control. These findings support its use as a potentially practical adjuvant in resource-limited settings

## Background

Orthopedic surgeries, particularly those involving the lower extremities, constitute a significant portion of surgical procedures worldwide. In the United States alone, an estimated 6.6 million orthopedic surgeries are performed annually, with a considerable proportion requiring lower limb interventions (Schreiber et al.,2019). In low- and middle-income countries (LMICs), especially in sub-Saharan Africa (SSA), the burden of orthopedic conditions is rising due to increasing rates of trauma, road traffic injuries, and degenerative conditions, often with limited access to adequate surgical care (Alayande *et al*., 2022). Studies estimate that 5.9 million people die annually from trauma-related causes worldwide, and a significant proportion of survivors suffer from long-term disabilities requiring orthopedic surgeries (Hwang, Jeong and Jo, 2023). Tanzania, like many SSA countries, faces a dual burden of trauma from road accidents and degenerative orthopedic conditions such as osteoarthritis, which places additional demand on surgical services (Blackman *et al*., 2024).

Among these surgical interventions, effective anesthesia is paramount to ensure patient comfort, minimize intraoperative pain, and support a seamless surgical experience (Komasawa, 2024). Subarachnoid Block (SAB), or spinal anesthesia, is widely used for lower extremity orthopedic surgeries due to its effective anesthesia, minimal cognitive impairment, and robust postoperative pain relief (Kamel, Ahmed and Sethi, 2022).

According to a retrospective review of 10,000 orthopedic surgeries, over 70% of lower limb procedures utilized SAB as the preferred anesthetic method (Liu *et al*., 2024). Additionally, SAB results in fewer cases of postoperative nausea and vomiting, reduced cardiorespiratory complications, and better post-operative pain coverage than general anesthesia, allowing patients to communicate effectively during surgery (Kaur *et al*., 2021). In resource-limited settings like Tanzania, SAB is often preferred due to its cost-effectiveness, safety profile, and reduced need for sophisticated monitoring equipment compared to general anesthesia (Dohlman, Kwikiriza and Ehie, 2020).

Intrathecal Bupivacaine, a long-acting local anesthetic, is frequently chosen for orthopedic surgeries due to its effective pain relief and strong sensory blockade (Sabertanha *et al*., 2023). However, studies have shown that the duration of analgesia provided by Bupivacaine alone can range from 2 to 3 hours (Nirmala *et al*., 2015). For longer surgical procedures or more extended postoperative recovery periods, the limited analgesic duration necessitates adjuncts to extend pain relief and manage the needs of patients effectively. In particular, opioid-based adjuvants have historically been employed, with notable success in enhancing analgesia (Abebe *et al*., 2022). However, systemic opioid-related side effects such as nausea (seen in 30-40% of patients), vomiting (15-25%), and respiratory depression have led to the exploration of alternatives (Swain *et al*., 2017). In SSA, the availability of opioids is often limited due to regulatory restrictions and supply chain issues, further driving the need for alternative methods of pain control (Cleary *et al*., 2013).

To minimize the reliance on systemic opioids and their associated side effects, there has been a growing interest in using adjuvants like dexamethasone with local anesthetics (Emelife *et al*., 2018). Dexamethasone, a potent corticosteroid with anti-inflammatory, analgesic, and antiemetic properties, has shown promise in enhancing the efficacy of local anesthetics (Jain and Dua, 2015). A randomized controlled trial involving 60 patients undergoing femur fracture surgeries demonstrated that adding dexamethasone to the anesthetic regimen significantly extended the analgesic duration by 35% compared to Bupivacaine alone (Persec *et al*., 2014). Similarly, research by (Bousabbeh *et al*., 2022) on 150 orthopedic patients found that dexamethasone improved both the onset and duration of sensory and motor blockade, reducing the need for intraoperative analgesics and improving patient satisfaction.

Dexamethasone’s ability to extend the duration of spinal anesthesia stems from its multiple mechanisms, including the inhibition of inflammatory mediators and reduction of neurogenic inflammation. Clinical trials have reported a prolongation of sensory blockade by 40-50% when dexamethasone is added to spinal anesthesia (Huynh, Marret and Bonnet, 2015). Notably, studies involving intravenous and perineural dexamethasone administration have shown that both methods are effective in extending the duration of peripheral nerve blocks, but the precise efficacy between the two routes remains under investigation.(Huynh, Marret and Bonnet, 2015) reviewed 12 clinical studies and found that intravenous dexamethasone administration extended the duration of nerve blocks by an average of 8 hours, compared to perineural administration, which extended the duration by 12 hours.

From an epidemiological perspective, the use of dexamethasone as an adjuvant for SAB in orthopedic surgeries not only improves the quality of anesthesia but may also contribute to better postoperative outcomes, including reduced opioid consumption (Jain and Dua, 2015). Opioid-related adverse events, such as respiratory depression and delayed bowel recovery, occur in 10-20% of postoperative patients (Gupta *et al*., 2018), and reducing their use can have significant public health implications. Furthermore, the enhanced recovery outcomes associated with dexamethasone, such as decreased inflammation and faster return to function, could lead to a shorter length of hospital stay, which is economically beneficial for both patients and healthcare systems. A meta-analysis by Elsharkawy et al. (2019), which included 14 randomized controlled trials involving 1,000 patients, showed that the addition of dexamethasone reduced the need for postoperative opioids by 25-30% and led to a 15% reduction in hospital stay duration for orthopedic patients (Kaiser *et al*., 2024).

In Tanzania, the benefits of dexamethasone in extending analgesia and reducing opioid use could be particularly impactful, given the limited availability of opioids and the high burden of trauma-related orthopedic surgeries. Furthermore, optimizing postoperative pain management through SAB and dexamethasone may improve patient outcomes and reduce the length of hospital stays, critical in resource-limited healthcare systems (Hyland *et al*., 2021).

Dexamethasone, when used with Bupivacaine in spinal anesthesia, has shown consistent efficacy in enhancing the quality and duration of anesthesia, reducing opioid requirements, and promoting faster recovery (Bousabbeh *et al*., 2022). As the global burden of orthopedic surgeries continues to grow, improving perioperative care through such adjuvants could have far-reaching benefits for patient outcomes and healthcare resource utilization (Beyranvand, Aryankhesal and Hashjin, 2019).

Despite the promising analgesic benefits of dexamethasone as an adjuvant in neuraxial anesthesia, concerns remain regarding the safety of intrathecal corticosteroid administration, particularly with preservative-containing formulations. Neurotoxicity has been recognized as an important consideration with intrathecal drug administration, as preservatives and excipients used in commercial preparations may potentially contribute to neural injury, although currently available evidence suggests that many commonly used preservatives may have relatively low neurotoxic potential. Existing clinical studies evaluating intrathecal dexamethasone have reported prolonged analgesic effects without major neurologic complications; however, long-term safety data remain limited and inconsistently reported. Neurotoxicity remains an important consideration in intrathecal drug administration, particularly with preservative-containing formulations, although available evidence suggests that many commercially used preservatives may have relatively low neurotoxic potential(1)

While dexamethasone as an adjuvant for spinal anesthesia has been studied in high-resource settings, data are not well established for lower resource settings. This study aims to further investigate the effectiveness of dexamethasone as an adjuvant to intrathecal Bupivacaine in Subarachnoid Block for orthopedic patients, specifically focusing on extending sensory blockade, reducing opioid consumption, and improving overall postoperative recovery. By conducting a randomized controlled trial, this study will contribute valuable insights into optimizing spinal anesthesia for orthopedic surgeries, particularly in resource-limited settings like SSA and Tanzania.

## Method

### Ethical statement

Ethical clearance for this study was granted by the Kilimanjaro Christian Medical University College Research Ethics and Review Committee (CRERC) in Moshi, Tanzania (**No. PG 87/2024**) and from the National Institute for Medical Research (NIMR) in Tanzania. This study was prospectively registered with the Pan African Clinical Trial Registry (PACTR), hosted by the South African Medical Research Council, under the identifier **PACTR202411679781198**.

### Study Design and Area

This study employed a double-blind, randomized clinical trial design to assess the effectiveness of dexamethasone as an adjunct to intrathecal bupivacaine compared to bupivacaine alone in lower limb orthopedic surgery patients undergoing spinal anesthesia. The study was conducted from January 2025 to May 2025. Participants were randomly assigned to either the Intervention Group (Dexamethasone *(Deexa®, Lincoln Pharmaceuticals Ltd, India)* and Bupivacaine group) or the Control Group (Bupivacaine and normal saline group) using simple randomization to ensure balanced allocation. Study focused on all adult patients (>18 years) scheduled for lower limb orthopedic surgeries under spinal anesthesia. KCMC is a tertiary, referral, and teaching hospital that receives patients from district and regional hospitals across the central and northern zones of Tanzania. The hospital has a 640-bed capacity and provides medical services to a population of approximately 15.7 million people. The anesthesia department is comprised of a dedicated team that includes 6 anesthesiologists, 22 anesthesia residents, and 16 nurse anesthetists. Most patients undergoing orthopedic surgeries at KCMC receive spinal anesthesia as the preferred anesthetic technique whenever indicated.

### Sample size

The sample size determination formula proposed by (Kane et al., 2024) and (Gupta *et al*., 2016)

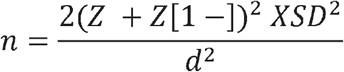

Whereby Level of significance = 5%,

Power = 90%

Zα = Z is constant set by convention according to accepted α error

Z (1-β) = Z is constant set by convention according to power of study which is calculated.

Zα = 1.96,

Z (1-β) = 1.28,

d - Effect size (From the previous study (Bani-hashem *et al*., 2011), which is equivalent to 30 minutes and a standard deviation of 10.

The sample size was derived by assuming an effect size of a mean difference of 9 minutes. While data from previous studies showed the effect size of the mean difference of around 30 minutes between arms, we assumed a lower effect size given the consideration that this trial will be a pragmatic trial done in clinical settings. There may be variabilities in techniques and experience that influence the outcomes. Also, there might be genetic variabilities between populations in previous studies and our population, so assuming a decreased effect size requires an increase the power of the study.

N = 43 patients in each arm

This gives us 96 participants with an inclusion of 10% for protocol deviations and late exclusion. N =96 participants, 48 patients in each arm.

### Randomization and Blinding

Participants were randomized using a computer-generated randomization sequence prepared by an independent investigator not involved in recruitment or outcome assessment. Group allocation was communicated only to the anesthetist responsible for preparation of the study drugs and administration of the spinal block. This anesthetist was not involved in postoperative data collection or outcome assessment.

### Data collection tools, methods and procedure

Data collection for this study employed a multifaceted approach integrating clinical assessments, medical record reviews, and patient monitoring to assess the impact of dexamethasone as an adjunct to intrathecal bupivacaine in orthopedic surgery patients who received spinal anesthesia. Initially, demographic information including age, gender, and ASA physical status classification was gathered from medical records (anesthesia chart records). Relevant medical history such as previous surgeries, allergies, chronic conditions, and current medications was also documented. Preoperatively, baseline assessments included vital signs, baseline pain scores, and evaluations of sensory and motor function. During surgery, details of anesthesia administration, including the type and dosage of bupivacaine with or without dexamethasone, were recorded. Postoperatively, objective assessments focused on the duration and quality of sensory and motor blocks using standardized scales at specified intervals. Pain intensity was assessed using a numerical rating scale, with data collected on time to the first analgesic request. Complications related to anesthesia were meticulously monitored. Trained personnel oversaw data collection procedures to ensure consistency, accuracy, and adherence to ethical guidelines throughout the study duration.

### Study Variables

#### Dependent variables

The effectiveness of dexamethasone as an adjuvant to intrathecal bupivacaine versus bupivacaine alone in orthopedic surgery was determined primarily by the total duration of sensory block, time to first post-operative analgesia and post-operative pain scores. The duration of the sensory block was defined as the time from the peak level of sensory block to the point when the patient first reported pain at the surgical site. The time to first postoperative analgesia was defined as the duration from the completion of the intrathecal injection to the first patient request or administration of rescue analgesia following surgery. Postoperative pain score was assessed using the Numerical Rating Scale (NRS), where patients rated their pain on a scale from 0 (no pain) to 10 (worst imaginable pain). Pain scores were recorded at regular intervals following surgery to evaluate the effectiveness of the spinal block and the need for rescue analgesia.

#### Independent variables

The study considered several predictors and independent variables related to the patients’ demographic, surgical, and health characteristics. Sex was treated as a binary variable coded as male or female, while age was measured as a continuous variable in years. Body Mass Index (BMI) was categorized into relevant groups based on standard BMI classifications. ASA classification (American Society of Anesthesiologists) was used to assess the physical status of patients prior to surgery. The type of surgery performed was recorded as a categorical variable based on the specific orthopedic procedure, and the length of surgery was measured as a continuous variable in minutes. Anesthetic provider was included as a categorical variable based on the provider’s qualification level. These predictors were analyzed to assess their influence on the study outcomes.

### Data Management and Analysis

Data were entered, cleaned, and double-checked for completeness and consistency. All data management and statistical analyses were performed using R Software for Statistical Computing version 4.5.1 in the integrated development environment Rstudio version 2026.0.1. Continuous variables were summarized using means and standard deviations, while categorical variables were summarized using frequencies and percentages. Comparisons between the characteristics and outcomes of control and intervention groups of patients were conducted using the Welch independent-sample t-test for continuous variables and Pearson’s chi-square test or Fisher’s exact test for categorical variables. To further evaluate the effect of the intervention, multivariable linear mixed-effects regression analyses were performed to estimate adjusted mean differences for continuous outcomes, controlling for potential confounders including age, gender, BMI, ASA class, type of surgery, and duration of surgery and accounting for the within-patients variability of repeated measures of the outcomes. A p-value of less than 0.05 was considered statistically significant

## Results

### Eligibility chart of the enrolled participants in the study

Among the 102 patients assessed for eligibility, 6 were excluded, 4 for not meeting the inclusion criteria and 2 who declined to participate. A total of 96 participants were therefore enrolled and randomized using a computer-generated sequence in a 1:1 ratio, with 50 allocated to the intervention group and 46 to the control group. All 96 randomized participants were included in the final analysis. (Fig. 1).

**Fig. 1.**
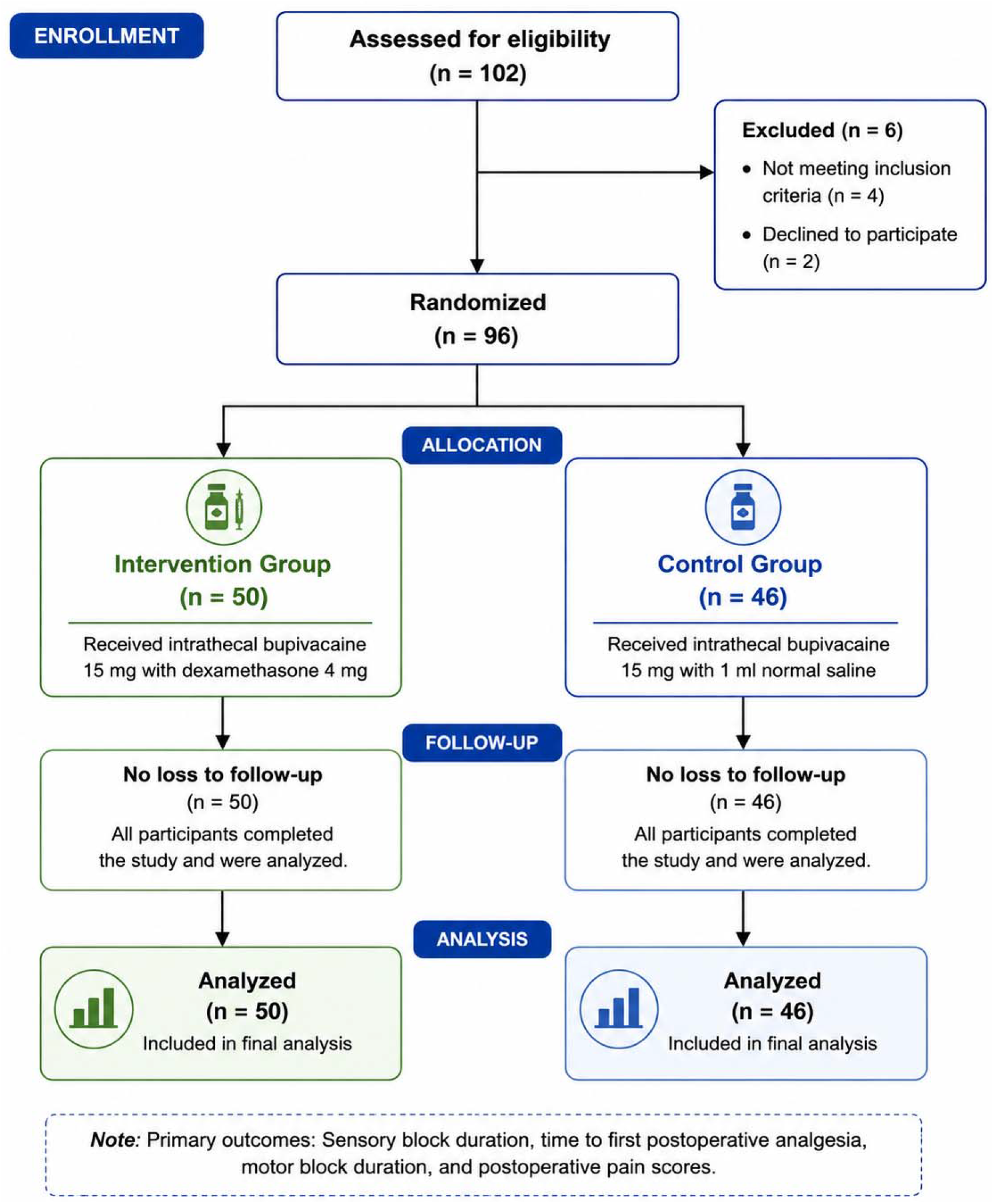

### Sociodemographic and clinical characteristics of the participants

The study included 96 orthopedic surgery patients with a mean age was 42 years (±16). Females predominated 56 (58%). The average BMI was 24.5 (±3.0) indicating a generally healthy weight range. The most common surgeries were thigh 38 (40%) and hip 25 (26%), followed by knee 15 (16%), ankle and foot 10 (10%), and leg surgeries 8 (8.3%). Intraoperative analgesia was rarely administered, with 85 (89%) receiving none. Opioids were given to 9 (9.4%), non-opioids to 1 (1%), and others to 1 (1%). Anxiolytics were also infrequently used, with benzodiazepines administered to 7 (7.3%). Most patients had no diabetes 88 (94%), while 6 (6.4%) had diabetes. Similarly, the majority had no hypertension 82 (86%), while 13 (14%) had hypertension. ASA class was mainly I 72 (75%), with 24 (25%) classified as ASA II. The mean duration of surgery was 125 minutes (±27). Spinal anesthesia was predominantly administered by residents 72 (76%), with nurse anesthetists covering 23 (24%). (**Table 1)**

**Table 1:** Comparison of Social and Clinical demographic characteristics between the two groups.

| Variable | Overall | Control | Intervention | P- value |
| --- | --- | --- | --- | --- |
| Age | 42 (16) | 43 (16) | 41 (15) | 0.5 |
| Gender |  |  |  | 0.11 |
| Male | 40 (42%) | 23 (50%) | 17 (34%) |  |
| Female | 56 (58%) | 23 (50%) | 33 (66%) |  |
| BMI | 24.5 (3.0) | 24.5 (3.2) | 24.4 (2.8) | >0.9 |
| Type of orthopedic surgery |  |  |  | 0.8 |
| Ankle and foot surgeries | 10 (10%) | 5 (11%) | 5 (10%) |  |
| Hip surgeries | 25 (26%) | 13 (28%) | 12 (24%) |  |
| Knee surgeries | 15 (16%) | 7 (15%) | 8 (16%) |  |
| leg surgeries | 8 (8.3%) | 5 (11%) | 3 (6.0%) |  |
| Thigh surgeries | 38 (40%) | 16 (35%) | 22 (44%) |  |
| Type of analgesia given intraoperatively |  |  |  | **0.010** |
| Non opioids | 1 (1.0%) | 1 (2.2%) | 0 (0%) |  |
| None | 85 (89%) | 37 (80%) | 48 (96%) |  |
| Opioids | 9 (9.4%) | 8 (17%) | 1 (2.0%) |  |
| Others | 1 (1.0%) | 0 (0%) | 1 (2.0%) |  |
| Anxiolytic given intraoperatively |  |  |  | **0.05** |
| Benzodiazepine | 7 (7.3%) | 6 (13%) | 1 (2.0%) |  |
| None | 89 (93%) | 40 (87%) | 49 (98%) |  |
| **Diabetes Mellitus** |  |  |  | 0.2 |
| No | 88 (94%) | 44 (98%) | 44 (90%) |  |
| Yes | 6 (6.4%) | 1 (2.2%) | 5 (10%) |  |
| **Hypertension** |  |  |  | >0.9 |
| No | 82 (86%) | 39 (87%) | 43 (86%) |  |
| Yes | 13 (14%) | 6 (13%) | 7 (14%) |  |
| **ASA class** |  |  |  | 0.8 |
| ASA 1 | 72 (75%) | 34 (74%) | 38 (76%) |  |
| ASA 2 | 24 (25%) | 12 (26%) | 12 (24%) |  |
| **Duration of surgery** | 125 (27) | 126 (27) | 124 (28) | 0.7 |
| **Who administered Spinal anesthesia** |  |  |  | 0.7 |
| Nurse Anesthetist | 23 (24%) | 10 (22%) | 13 (26%) |  |
| Resident | 72 (76%) | 35 (78%) | 37 (74%) |  |

<sup>1</sup>Mean (SD); n (%) <sup>2</sup>Welch Two Sample t-test; Pearson's Chi-squared test; Fisher's exact test

### Comparison of Social and Clinical demographic characteristics between the two groups

The intervention and control groups were comparable in terms of age (p = 0.5), sex distribution (p = 0.11), BMI (p > 0.9), ASA class (p = 0.8), comorbidities including diabetes mellitus (p = 0.2) and hypertension (p > 0.9), and types of orthopedic surgery performed (p = 0.8). There was also no significant difference in the duration of surgery (p = 0.7) or the cadre of anesthesia provider (p = 0.7). However, statistically significant differences were observed in intraoperative management. The control group had a higher use of opioids (p = 0.010) and benzodiazepine anxiolytics (p = 0.05), suggesting greater intraoperative analgesic and anxiolytic requirements in the absence of dexamethasone (**Table 1**)

### Comparison of Sensory Block Duration, Time to First Analgesia, and Postoperative Pain Scores Between Control and Intervention Groups

The intervention group demonstrated a significantly prolonged duration of sensory blockade compared to controls (231 ± 6 vs. 156 ± 9 minutes; p < 0.001), indicating enhanced anesthetic efficacy. Similarly, the mean time to first postoperative analgesia was significantly extended in the intervention group (252 ± 7 minutes) compared to the control group (181 ± 7 minutes; p < 0.001), reflecting improved postoperative pain control. In addition, the duration of motor blockade was significantly longer in the intervention group (184 ± 7 vs. 130 ± 5 minutes; p < 0.001).

Analysis of postoperative pain scores revealed significantly lower pain intensity in the intervention group at 1 hour (1.06 vs. 6.63; p < 0.001) and 2 hours (2.49 vs. 4.46; p < 0.001), as well as at 12 hours (3.10 vs. 3.40; p = 0.014). However, no statistically significant differences were observed at 3 hours (p = 0.06), 4 hours (p = 0.2), and 6 hours (p = 0.3). (**Table 2**)

**Table 2:**
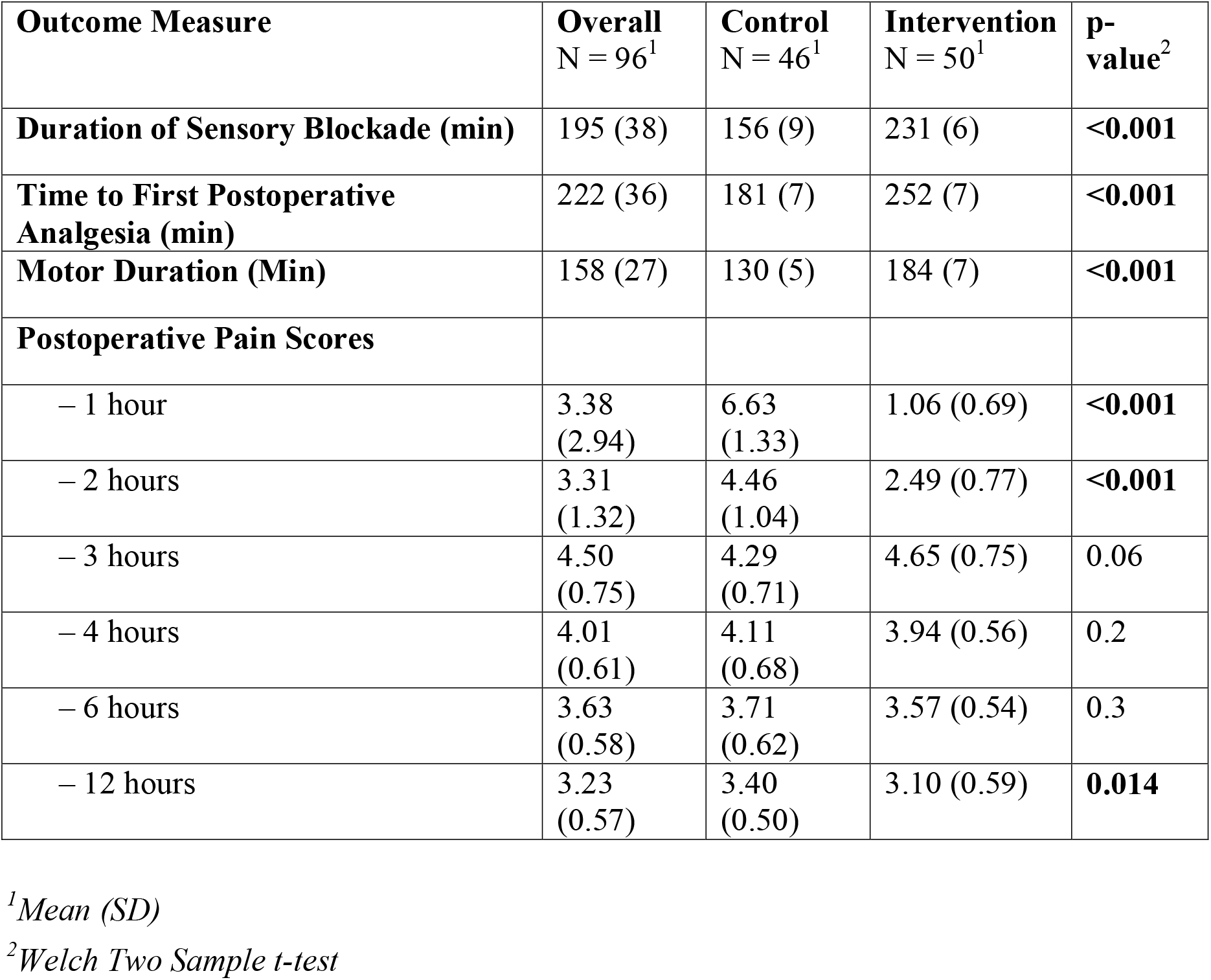
Comparison of Sensory Block Duration, Time to First Analgesia, and Postoperative Pain Scores Between Control and Intervention Groups.

### Adjusted Treatment Effect for the Intervention arm compared to the treatment arm

After adjusting for age, gender, BMI, ASA class, surgery type, and duration, the intervention group demonstrated significantly improved block and analgesic outcomes compared with controls. Sensory block lasted on average 75.11 minutes longer (95% CI: 71.92–78.29, *p* < 0.001), while motor block was extended by 53.42 minutes (95% CI: 50.99–55.85, *p* < 0.001). The time to first analgesia was delayed by 71.89 minutes (95% CI: 68.91–74.86, *p* < 0.001). Pain scores, adjusted for repeated measures, were 1.29 points lower (95% CI: −1.52 to −1.05, *p* < 0.001). Overall, the intervention significantly prolonged sensory and motor block, postponed analgesia requirements, and reduced postoperative pain, while hemodynamic parameters remained comparable between groups (**Table 3**).

**Table 3:**
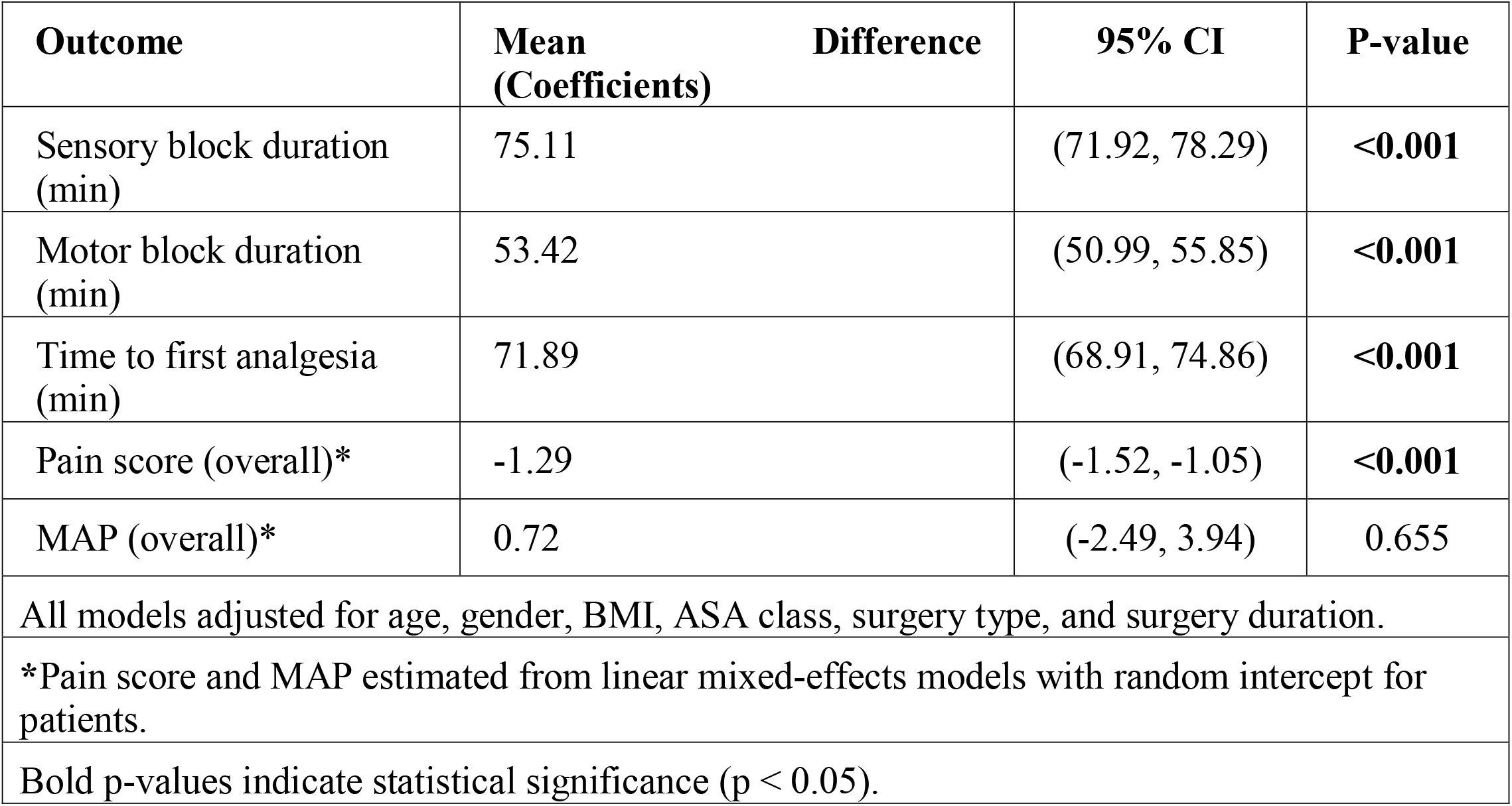
Adjusted Treatment Effect for the Intervention arm compared to the treatment arm.

## Discussion

This study evaluated the effectiveness of intrathecal dexamethasone as an adjuvant to bupivacaine in prolonging sensory block duration, delaying the time to first postoperative analgesia, and reducing postoperative pain in patients undergoing orthopedic surgery under spinal anesthesia. The results demonstrated significant benefits of dexamethasone as an adjunct, aligning with existing literature and providing further insight into its applicability in low-resource settings such as ours.

The findings of this study showed that patients in the intervention group had a significantly longer duration of sensory block compared to the control group (231 ± 6 vs. 156 ± 9 minutes; p < 0.001). This substantial difference supports the hypothesis that intrathecal dexamethasone prolongs sensory blockade when combined with bupivacaine. These findings are consistent with other studies. For example, a study conducted in India by (2) reported the overall duration of sensory block was prolonged significantly in the dexamethasone group (311.43 ± 13 minutes) when compared to other groups, fentanyl group (197.86 ± 12 minutes) and bupivacaine alone with normal saline group (115.29 ± 14.6 minutes). Similarly, (3) also found significantly longer block durations in the dexamethasone group (119±10.69 minutes) compared to the control group (89.44±8.37 minutes) in spinal anesthesia for orthopedic surgery, the analgesic and block-prolonging effects of dexamethasone may be explained through several proposed mechanisms. Dexamethasone acts on glucocorticoid receptors, leading to modulation of nociceptive signal transmission and suppression of inflammatory mediator release around neural tissues. In addition, corticosteroids are thought to reduce ectopic neuronal discharge and suppress transmission in unmyelinated C-fibers, thereby decreasing pain propagation. Its potent anti-inflammatory effects may also reduce local inflammatory responses and neurogenic inflammation, contributing to prolonged sensory blockade and improved postoperative analgesia.

The extended duration of sensory block and analgesia may allow longer procedures to be completed under spinal anesthesia, reducing the need for intraoperative conversion to general anesthesia. This is especially beneficial in low-resource settings where general anesthesia may pose increased risks due to limited monitoring and airway equipment. Moreover, in environments where advanced pain management options such as epidural analgesia or peripheral nerve blocks are unavailable, and where adjuvants like dexmedetomidine or opioids face cost or supply chain limitations, dexamethasone offers a practical, cost-effective alternative.

In terms of postoperative analgesia, the intervention group demonstrated a significantly delayed need for analgesics (mean time to first postoperative analgesia: 252 ± 7 minutes vs. 181 ± 7 minutes in controls; p < 0.001). These findings were further supported by adjusted analysis, which showed that dexamethasone prolonged the time to first analgesic request by approximately 71.9 minutes (95% CI: 68.91–74.86; p < 0.001). This is in line with previous research indicating that dexamethasone not only extends the duration of spinal anesthesia but also enhances postoperative analgesia. The study conducted in Tunisia by (4) reported similar findings. Similarly a study done in Iran by (5) also showed that the average time to first analgesic request in the case group was 345.83±0.4 minutes (dexamethasone group) and 251.52±1.8 minutes in the Bupivacaine alone group which is also consistent with the findings from this study.

Pain scores postoperatively were also consistently lower in the intervention group, particularly during the early postoperative period. Statistically significant reductions in pain were observed at 1 hour (1.06 vs. 6.63; p < 0.001) and 2 hours (2.49 vs. 4.46; p < 0.001), However, no significant differences were noted at 3, 4, and 6 hours postoperatively. These findings suggest that dexamethasone provides superior early postoperative analgesia. The adjusted analysis further confirmed an overall reduction in pain scores by 1.29 points (95% CI: −1.52 to −1.05; p < 0.001), reinforcing the analgesic benefit of dexamethasone. These findings are consistent with studies by (6) and (2), both of which noted improved postoperative pain control with the use of dexamethasone. Although patient satisfaction was not directly measured in this study, the marked difference in early postoperative pain scores suggests that patients in the dexamethasone group may have experienced greater comfort during recovery. It is plausible that these lower pain levels could translate into improved patient satisfaction, reduced opioid consumption, and enhanced recovery. While these outcomes were beyond the scope of the present study, they represent important clinical implications and warrant further investigation in future research(7).

Notably, differences were also observed in intraoperative medication requirements between the two groups. Patients in the control group were significantly more likely to require intraoperative opioids (17% vs. 2%; p = 0.010) and benzodiazepines (13% vs. 2%; p = 0.05) compared to those in the intervention group. These findings suggest that the addition of dexamethasone improved intraoperative analgesia and patient comfort, thereby reducing the need for supplemental medications. This further supports the role of dexamethasone as an effective adjuvant not only in prolonging block duration but also in enhancing intraoperative conditions.

The intervention group also demonstrated a prolonged duration of motor blockade compared to the control group (184 ± 7 vs. 130 ± 5 minutes; p < 0.001), with an adjusted mean difference of 53.42 minutes (95% CI: 50.99–55.85; p < 0.001). While this reflects an extended pharmacologic effect of the anesthetic regimen, its clinical relevance should be interpreted cautiously. Although the prolongation of motor blockade reached statistical significance, the magnitude of the difference (∼50 minutes) is relatively small and unlikely to be clinically meaningful, as early mobilization in most lower limb procedures is not typically undertaken within the immediate postoperative period. Nevertheless, prolonged motor block may potentially delay early mobilization and rehabilitation, particularly in enhanced recovery after surgery (ERAS) pathways, and this should be balanced against the analgesic benefits of dexamethasone.

### Strengths and Limitations of the Study

This study had several strengths. Firstly, it utilized objective and standardized methods to assess outcomes such as duration of block, time to analgesia, and pain scores using the NRS at multiple fixed intervals. Secondly, the double-blind design minimized bias during data collection and outcome assessment. Thirdly, this is one of the few studies to assess intrathecal dexamethasone use in the context of a low-resource orthopedic surgery setting, providing valuable contextual insight for clinical practice at KCMC and similar centers.

However, the study had some limitations. The study was conducted at a single center, which may limit the generalizability of the findings to other institutions or surgical contexts. While the study adjusted for several confounders, residual confounding from unmeasured variables (such as surgical technique variability or Surgeon’s experience) cannot be completely excluded. This study did not specifically evaluate safety outcomes such as neurologic complications, urinary retention, infection, or other potential steroid-related adverse effects following intrathecal dexamethasone administration. Therefore, conclusions regarding the safety profile of intrathecal dexamethasone cannot be definitively drawn from this study. Another limitation is that preservative-free dexamethasone was unavailable during the study period. While no neurologic adverse events were identified clinically, long-term safety outcomes related to intrathecal administration were not formally evaluated.

## Conclusion

Intrathecal dexamethasone was associated with prolonged sensory block, delayed postoperative analgesic requirement, and improved early postoperative pain scores in this study population.

## Data Availability

All data produced in the present study are available upon reasonable request to the authors

## Authors Contributions

**KF, BK** contributed to study design, patient recruitment, and data collection. **WN, ES**, and **LVM** led the data analysis and interpretation, with daily involvement throughout the study. **GG** supported clinical coordination and quality assurance during data collection. **AL, VA, AD, HS, FM, MM** provided methodological guidance and critically revised the manuscript. **NC and BV** supervised the research process, contributed to the study conception, and provided senior oversight.

## Conflict of Interest

All authors have no conflict of interest to disclose

## Funding Statement

This study did not receive any funding

## Notes

### Competing Interest Statement

The authors have declared no competing interest.

### Clinical Trial

PACTR202411679781198

### Author Declarations

Ethical clearance for this study was granted by the Kilimanjaro Christian Medical University College Research Ethics and Review Committee (CRERC) in Moshi, Tanzania (No. PG 87/2024) and from the National Institute for Medical Research (NIMR) in Tanzania. This study was prospectively registered with the Pan African Clinical Trial Registry (PACTR), hosted by the South African Medical Research Council, under the identifier PACTR202411679781198.

